# Early analysis of a potential link between viral load and the N501Y mutation in the SARS-COV-2 spike protein

**DOI:** 10.1101/2021.01.12.20249080

**Authors:** Tanya Golubchik, Katrina A. Lythgoe, Matthew Hall, Luca Ferretti, Helen R. Fryer, George MacIntyre-Cockett, Mariateresa de Cesare, Amy Trebes, Paolo Piazza, David Buck, John A. Todd, The COVID-19 Genomics UK (COG-UK) consortium, Christophe Fraser, David Bonsall

## Abstract

A new variant of SARS-CoV-2 has emerged which is increasing in frequency, primarily in the South East of England (lineage B.1.1.7 (*1*); VUI-202012/01). One potential hypothesis is that infection with the new variant results in higher viral loads, which in turn may make the virus more transmissible. We found higher (sequence derived) viral loads in samples from individuals infected with the new variant with median inferred viral loads were three-fold higher in individuals with the new variant. Most of the new variants were sampled in Kent and Greater London. We observed higher viral loads in Kent compared to Greater London for both the new variant and other circulating lineages. Outside Greater London, the variant has higher viral loads, whereas within Greater London, the new variant does not have significantly higher viral loads compared to other circulating lineages. Higher variant viral loads outside Greater London could be due to demographic effects, such as a faster variant growth rate compared to other lineages or concentration in particular age-groups. However, our analysis does not exclude a causal link between infection with the new variant and higher viral loads. This is a preliminary analysis and further work is needed to investigate any potential causal link between infection with this new variant and higher viral loads, and whether this results in higher transmissibility, severity of infection, or affects relative rates of symptomatic and asymptomatic infection

**Document Description and Purpose:** This is an updated report submitted to NERVTAG in December 2020 as part of urgent investigations into the new variant of SARS-COV-2 (VUI-202012/01). It makes full use of (and is restricted to) all sequence data and associated metadata available to us at the time this original report was submitted and remains provisional. Under normal circumstances more genomes and metadata would be obtained and included before making this report public. We will update this preprint when more genomes and metadata are available and before submitting for peer review.

## Background

On 14 December 2020 a new variant of SARS-CoV-2 circulating in the UK was reported (*2, 3*), characterised by the N501Y mutation in the receptor binding domain (RBD) of Spike, the ΔH69/V70 deletion, and numerous other mutations (*1*). The rise in frequency of this variant is associated with a sharp increase in reported cases in the South East of England, raising concerns that the variant could be more transmissible. We performed a rapid analysis to investigate whether the new variant is associated with higher viral loads, since higher viral loads may indicate increased transmissibility.

## Methods

As members of the COG-UK consortium (https://www.cogconsortium.uk/), we sequenced RT-QPCR SARS-CoV-2 positive samples originating from four UK Lighthouse laboratories, which provide Pillar 2 COVID-19 testing services. The samples were sequenced using veSEQ, our quantitative sequencing approach for which the number of unique mapped reads is correlated with, and thus can be used as a proxy for, viral load. For a full description of the sequencing protocol see (*4, 5*).

We used log_10_ (mapped reads) as a proxy for viral load (see fig S1 in (*5*)). Comparisons between distributions of log_10_ (mapped reads) were made using Welsh t-test (two-tailed), with p-values combined using Stouffer’s method where appropriate. We also performed a multivariate logistic regression analysis.

### Number of unique mapped reads is negatively correlated with Ct value

Given the known negative correlation between viral load and cycle threshold (Ct) values (*6*) obtained during PCR testing (*7*), we first confirmed a strong negative correlation between log_10_(unique mapped reads) and Ct values for samples that we sequenced from Lighthouse laboratories (linear regression, r^2^=0.43, *p*<<0.001, Fig. 1).

**Figure 1.**
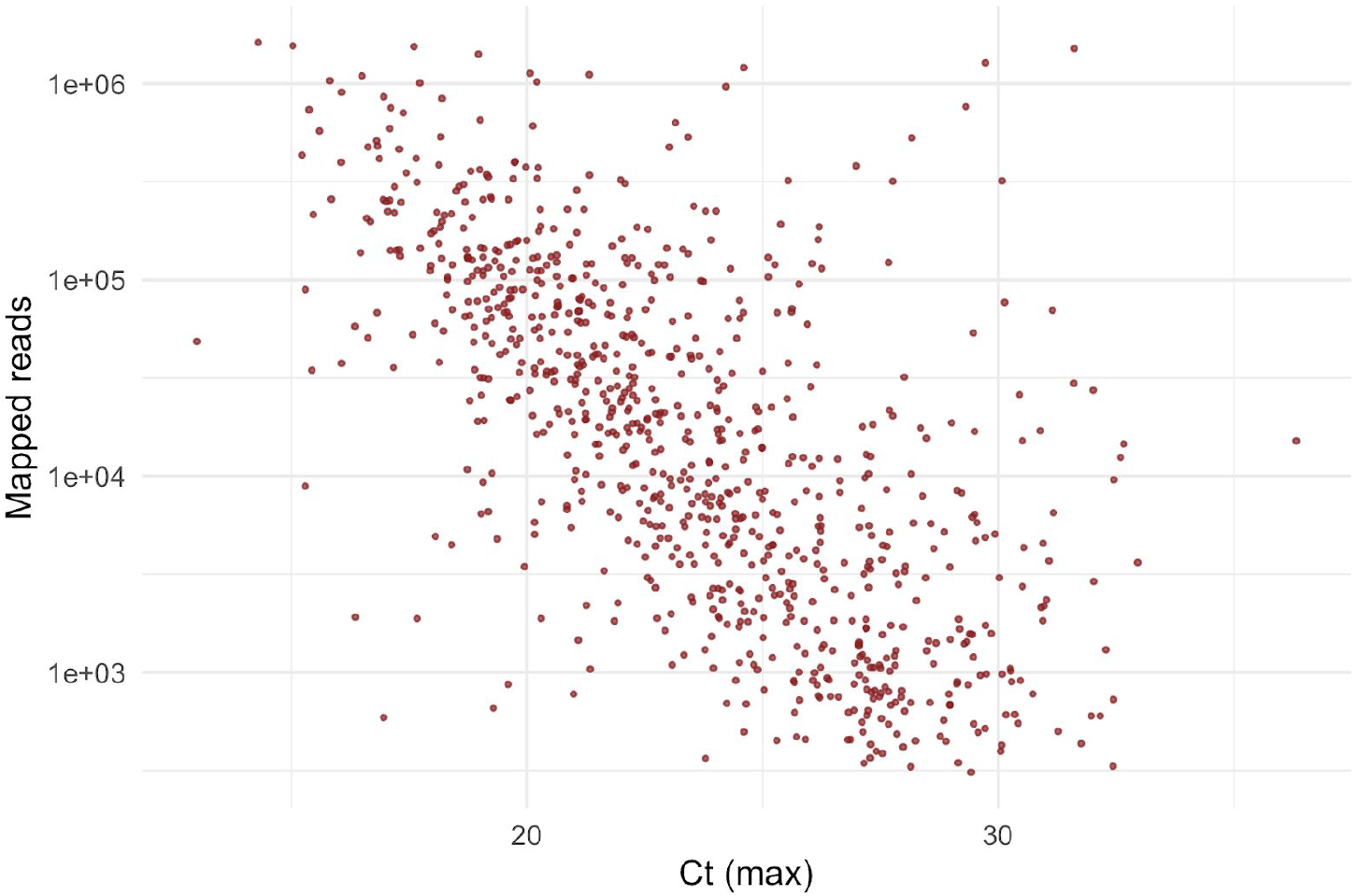
A strong negative correlation between Ct value and log10(number of mapped reads). Number of uniquely mapped reads per sample can be used as a proxy for viral load. The Ct value shown is the maximum Ct value obtained from Majora (the COG database) from retrospective data for all Lighthouse laboratories that supply Ct data; log10 of uniquely mapped (deduplicated) reads obtained with veSEQ platform correlates well with Ct. This does not include samples in this report since Ct values were not yet available.

**Figure 1:**
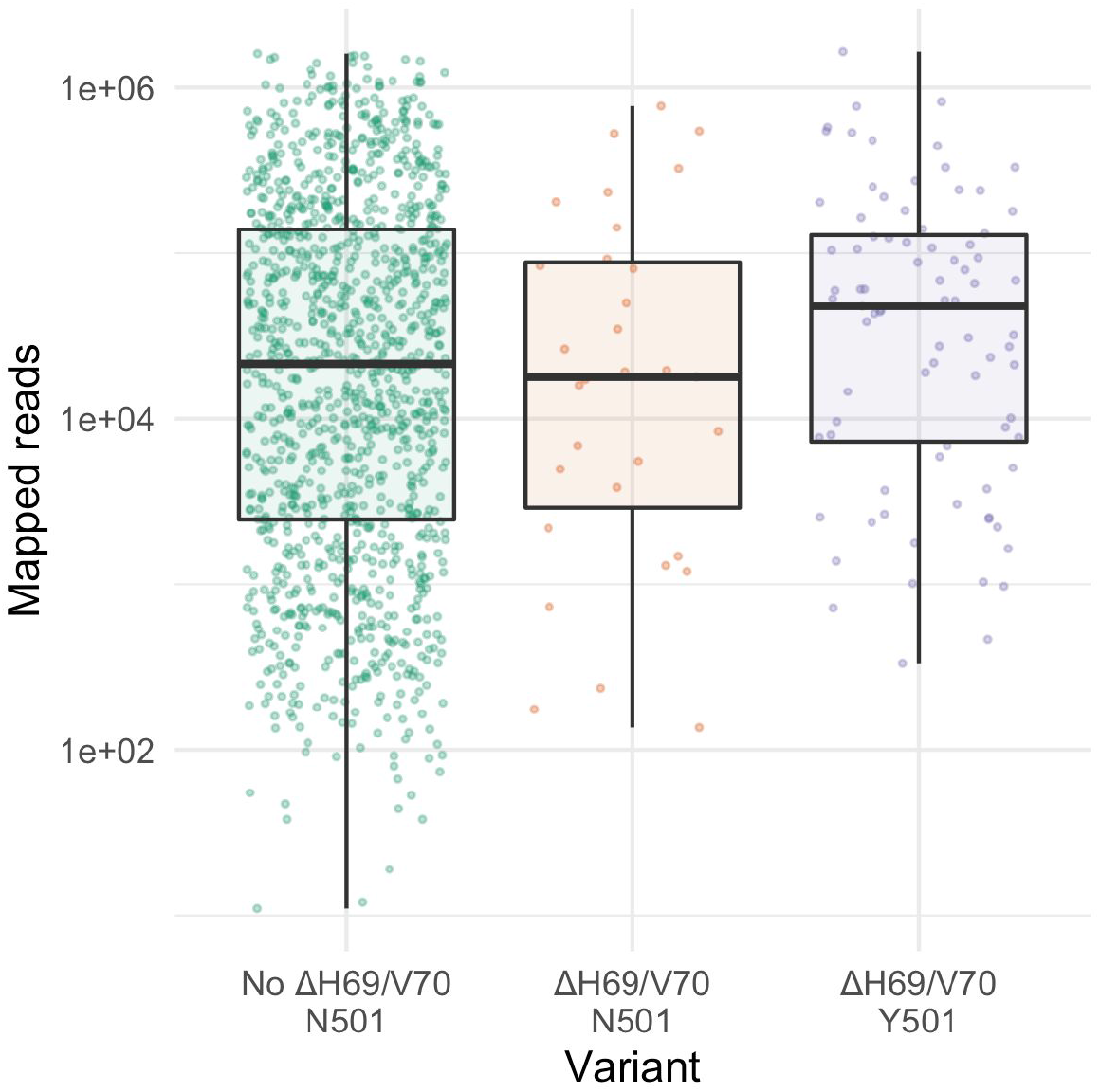
Higher numbers of mapped reads in samples with the Y501 variant. Box and scatter plots of unique mapped reads, stratified according to variant. Points within each batch are jittered to aid visualisation. Horizontal lines in boxplots represent the median and the interquartile range. The Y501 variant has a higher number of mapped reads, whereas the ΔH69/V70 deletion only has a higher number of mapped reads in the presence of the N501Y mutation.

### The new variant is associated with higher viral loads

The N501Y mutation is strongly linked with other mutations characterising the new variant (VUI-202012/01) in our dataset, including the ΔH69/V70 deletion, and therefore we used Y501 as a marker of the new variant. The ΔH69/V70 deletion alone is not a specific marker of VUI-202012/01 in our data, while lineage B.1.1.70, which is currently present in Wales and in some cases carries Y501 but never the deletion, was not present in our data.

We identified 88 samples that produced consensus sequences with the Y501 variant. All variant samples were taken between 31 Oct 2020 and 13 Nov 2020, and therefore we only considered samples (Y501 and N501) taken during this period, since Ct values have been shown to vary by calendar time (*7*).

When comparing the number of unique mapped reads in the Y501 variant samples (median log_10_(reads)=4.64, *N*=88) with that in the to N501 samples (median log_10_(reads)=4.16, *N*=1299), we found higher counts in the former (Welch *t*-Test *p*=0.014; Fig. 1). This is equivalent to around 3-fold higher median viral loads in the Y501 variant samples compared to N501 samples.

This result remained significant when we controlled for batch (Fig. 3a, *p*=0.011, combined *p*-value via Stouffer’s method), but not Lighthouse laboratory (Fig. 3b, *p*=0.052). The correlation between the new variant and viral load is also associated with a relative paucity of samples with lower (<10^3^) mapped reads among Y501 samples (Fig. 1, *p=*0.0053, chi-squared test; 10^3^ logged mapped reads is equivalent to a viral load of ∼10^4^ copies per reaction, max Ct∼28). When comparing samples with just the ΔH69/V70 deletion (without the Y501 variant) to samples without the deletion, we did not find a significant difference in log_10_(reads) (*p*=0.86; controlling for batch *p*=0.56, and for Lighthouse lab *p*=0.54) (Fig. 1).

**Figure 3.**
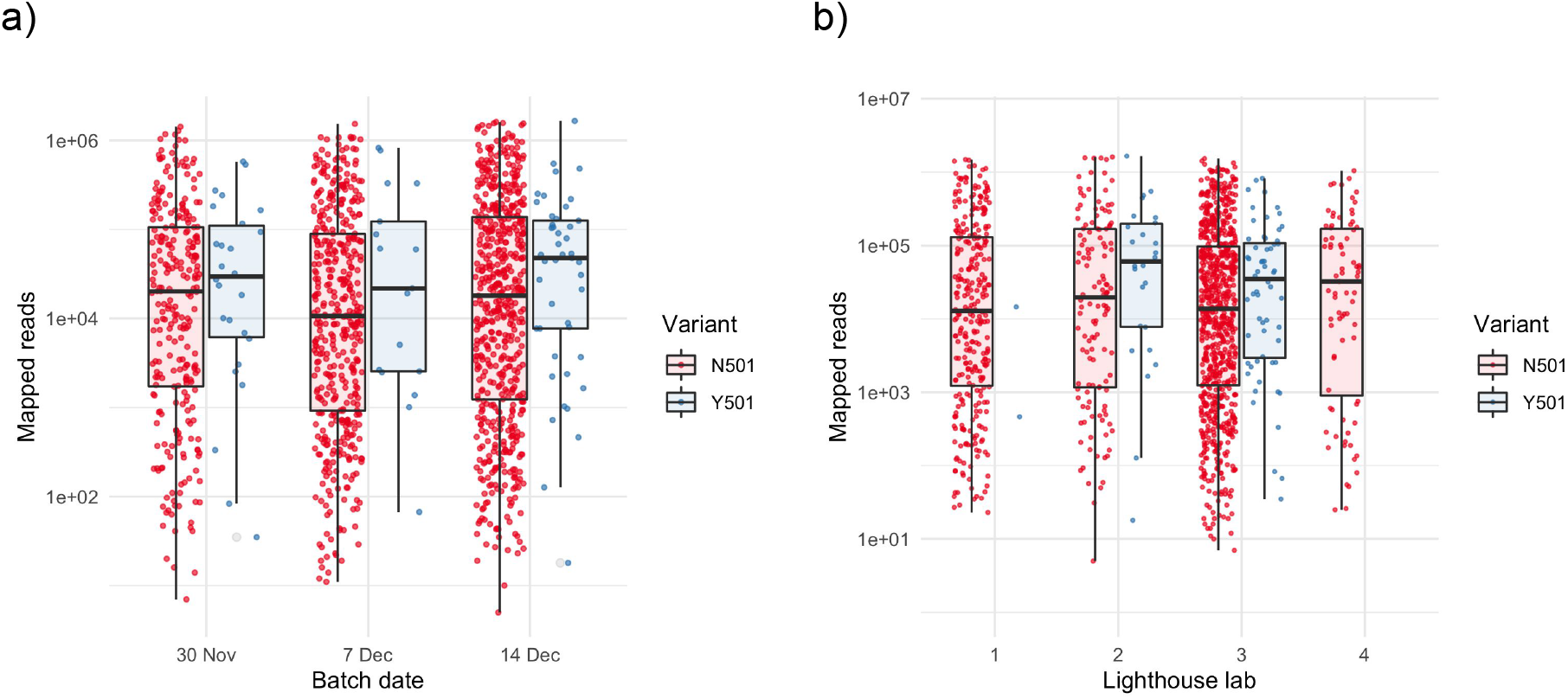
Higher numbers of mapped reads in samples exhibiting the Y501 variant. Box and scatter plots of unique mapped reads, stratified by (a) batch date and (b) anonymised Lighthouse lab. There is no significant difference among batches or Lighthouse labs for N501 samples (p>0.1 for all pairwise comparisons). Points within each batch are jittered to aid visualisation.Horizontal lines in boxplots represent the median and the interquartile range.

### Viral loads differ by sampling location

To test whether the difference in viral loads for samples with the new variant could in part be explained by geographic effects, we considered the sampling location (adm2 district) where this information was available. Of the 88 Y501 variants sampled, 24 were in Greater London, 46 in Kent, and in lower numbers (*N*=1-5) in other areas (Bristol, Essex, Hampshire, Leicestershire, Norfolk, Surrey and West Sussex). Regardless of variant presence, all samples from Greater London had significantly lower viral loads than those from other locations (*p*=0.0016, Welch’s *t*-test), and the association between Y501 and higher viral load was not significant in this region (p=0.91; Fig. 4). Outside Greater London, viral loads for Y501 were significantly higher than for N501 (*p*=0.0068). Within Kent, the location with the greatest number of Y501 samples, Y501 viral loads were not significantly higher than N501 viral loads (*p*=0.089). These results indicate a correlation between infection with the new variant (VUI-202012/01) and (inferred) viral load outside Greater London, although we are currently underpowered to draw firm conclusions. The lack of association within Greater London could be due to lack of power, or to demographic or epidemiological differences in London compared with the other locations.

**Figure 4.**
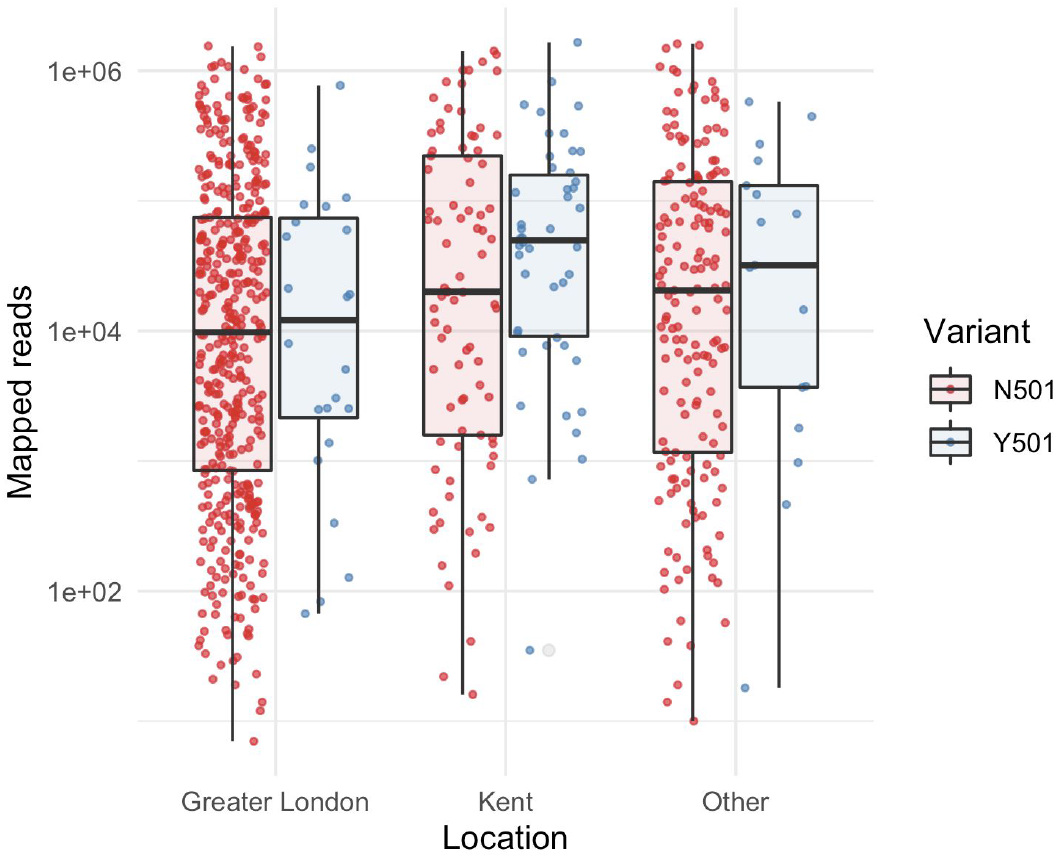
Number of mapped reads varies by sampling location. Box and scatter plots of unique mapped reads, stratified by sampling location. Points within each batch are jittered to aid visualisation. Horizontal lines in boxplots represent the median and the interquartile range. Only sampling locations with at least one Y501 sample were included.

In a multivariate logistic regression analysis for variables associated with higher viral load (Table 1), the Y501 variant was associated with a fivefold increase in odds of >=10^3^ mapped reads (*p*=0.036). The fitted model with interaction terms suggest a much smaller effect of the variant outside Kent, with the total odds increase reduced to 1.75 for Greater London and 1.24 for other regions, but the interaction term coefficients were not statistically significant (*p*=0.27 and *p*=0.16, respectively). Thus, if the association of the variant with a paucity of low viral load samples is stronger in Kent compared to other areas (e.g. due to epidemiological, demographic, or sampling differences), we lack the necessary power to demonstrate it. No other variables showed evidence of an association.

**Table 1.** Logistic regression analysis, identifying variables associated with >=10^3^ mapped reads.

|  | Odds ratio (95% CI) |
| --- | --- |
| Y105 | 5.078* (1.111, 23.218) |
| Region (reference: Kent) |  |
| Greater London | 0.675 (0.371, 1.227) |
| Other | 0.869 (0.476, 1.588) |
| Lighthouse lab (reference: 1) |  |
| 2 | 1.148 (0.703, 1.874) |
| 3 | 1.183 (0.610, 2.296) |
| 4 | 0.906 (0.512, 1.605) |
| Batch date (reference: 30 Nov) |  |
| 7 Dec | 0.822 (0.575, 1.173) |
| 14 Dec | 0.955 (0.502, 1.816) |
| Interaction: Y105 × Greater London | 0.345 (0.053, 2.248) |
| Interaction: Y105 × Other region | 0.245 (0.034, 1.771) |
| Observations | 1,376 |
| Note: *p<0.05; **p<0.01; ***p<0.001 |  |

Odds ratios for each variable and 95% confidence intervals for those ratios are presented. Lighthouse labs are anonymised as in Fig. 2.

### Caveats and Limitations

This is a preliminary analysis, and other factors could explain the (inferred) higher viral loads in samples with the new variant (VUI-202012/01), in addition to a working hypothesis that there is a causal effect of the new variant on within-host virus abundance. Whether the correlation is causative (infections with the new variant have higher viral loads) or correlative (e.g. due to epidemiological dynamics, demographics of individuals infected with the new variant, and/or sampling) warrants further study.

Individuals contributing samples in this analysis were tested as part of the test and trace program, which is primarily aimed towards individuals seeking a test following the onset of symptoms. We observed a broad spectrum of viral loads among the samples we sequenced. Given known associations between lower viral loads and later infection (*8*), and higher viral loads at the onset of symptoms, this suggests our full dataset consists of individuals in both early and late stages of symptomatic infection. Whilst we do not *a priori* expect there to be a systematic difference in the timing of sampling relative to infection, in an exponentially growing population the expectation is to sample relatively more people early in infection (*9*). Whether or not early sampling-bias supports an effect on inferred viral loads will depend on the relative epidemiological dynamics of the new and other variants. If, for example, VUI-202012/01 is growing faster, this could result in a bias for it to be sampled earlier. This is consistent with the relative paucity of VUI-202012/01 samples with low viral load.

In addition, VUI-202012/01 might be circulating within particular demographics (e.g. age groups) that tend to have higher viral loads when sampled. This may explain the apparently different patterns in Greater London and elsewhere. Focussed transmission within a particular demographic group is also more likely during the early stages of epidemic growth of a given lineage, before it disperses into the wider population. We were unable to test these hypotheses as we did not have demographic data relating to the sampled individuals with the new variant. We also cannot rule out other additional confounding effects and recommend that such effects are investigated further.

### Future prospects

A number of processess could have caused the rapid growth of the new variant (VUI-202012/01), including founder effects, or biological mechanisms that increase its transmissibility. Higher viral loads are one such potential mechanism: Transmissibility of viruses is understood to be higher in individuals who exhibit higher viral loads (*10*) and in HIV viral load is partly determined by virus genotype (*11*). Our observation of higher inferred viral loads in individuals infected with the new variant suggests that increased transmissibility of the new variant is plausible, but important caveats remain.

We recommend further investigations to evaluate this hypothesis. We note that we have used Y501 as a marker for the new variant; a large number of other mutations also characterise this new variant lineage (*1*), and therefore Y501 *per se* might not be causing the effect (if there is one). We also note that higher viral loads can be associated with higher levels of viral virulence, and therefore links between the new variant and the severity of infection should be monitored carefully (*12*).

Whether or not observed higher viral loads associated with this variant are a direct cause of infection with the variant, a consequence of faster epidemic growth, or linked to particular demographics, our data are consistent with rapid growth of this specific lineage.

## Data Availability

All genomic data has been made publicly available as part of the COVID-19 Genomics UK (COG-UK) Consortium via GISAID and via the European Nucleotide Archive (ENA) study PRJEB37886.

## Acknowledgments

We are grateful to Lorne Lornie, Angie Green, The Oxford Genomics Centre and The Wellcome Centre for Human Genetics, for all their support in generating the data for this study.

## Ethics

The COVID-19 Genomics UK (COG-UK) consortium study protocol was approved by the Public Health England Research Ethics and Governance Group (reference: R&D NR0195) on the 8th of April 2020. The decision was communicated to Professor Sharon Peacock by Dr Elizabeth Coates, Head of Research Governance, PHE Research Support and Governance Office.

## Appendix

### COG-UK Full list of consortium names and affiliations

#### Funding acquisition, leadership, supervision, metadata curation, project administration, samples, logistics, Sequencing, analysis, and Software and analysis tools

Thomas R Connor ^33, 34^, and Nicholas J Loman ^15^.

##### Leadership, supervision, sequencing, analysis, funding acquisition, metadata curation, project administration, samples, logistics, and visualisation

Samuel C Robson ^68^.

##### Leadership, supervision, project administration, visualisation, samples, logistics, metadata curation and software and analysis tools

Tanya Golubchik ^27^.

##### Leadership, supervision, metadata curation, project administration, samples, logistics sequencing and analysis

M. Estee Torok ^8, 10^.

##### Project administration, metadata curation, samples, logistics, sequencing, analysis, and software and analysis tools

William L Hamilton ^8, 10^.

##### Leadership, supervision, samples logistics, project administration, funding acquisition sequencing and analysis

David Bonsall ^27^.

##### Leadership and supervision, sequencing, analysis, funding acquisition, visualisation and software and analysis tools

Ali R Awan ^74^.

##### Leadership and supervision, funding acquisition, sequencing, analysis, metadata curation, samples and logistics

Sally Corden^33^.

##### Leadership supervision, sequencing analysis, samples, logistics, and metadata curation

Ian Goodfellow ^11^.

##### Leadership, supervision, sequencing, analysis, samples, logistics, and Project administration

Darren L Smith ^60, 61^.

##### Project administration, metadata curation, samples, logistics, sequencing and analysis

Martin D Curran ^14^, and Surendra Parmar ^14^.

##### Samples, logistics, metadata curation, project administration sequencing and analysis

James G Shepherd ^21^.

##### Sequencing, analysis, project administration, metadata curation and software and analysis tools

Matthew D Parker ^38^ and Dinesh Aggarwal ^1, 2, 3^.

##### Leadership, supervision, funding acquisition, samples, logistics, and metadata curation

Catherine Moore ^33^.

##### Leadership, supervision, metadata curation, samples, logistics, sequencing and analysis

Derek J Fairley^6, 88^, Matthew W Loose ^54^, and Joanne Watkins ^33^.

##### Metadata curation, sequencing, analysis, leadership, supervision and software and analysis tools

Matthew Bull ^33^, and Sam Nicholls ^15^.

##### Leadership, supervision, visualisation, sequencing, analysis and software and analysis tools

David M Aanensen ^1, 30^.

##### Sequencing, analysis, samples, logistics, metadata curation, and visualisation

Sharon Glaysher ^70^.

##### Metadata curation, sequencing, analysis, visualisation, software and analysis tools

Matthew Bashton ^60^, and Nicole Pacchiarini ^33^.

##### Sequencing, analysis, visualisation, metadata curation, and software and analysis tools: Anthony P Underwood ^1, 30^

##### Funding acquisition, leadership, supervision and project administration

Thushan I de Silva ^38^, and Dennis Wang ^38^.

##### Project administration, samples, logistics, leadership and supervision

Monique Andersson^28^, Anoop J Chauhan ^70^, Mariateresa de Cesare ^26^, Catherine Ludden ^1,3^, and Tabitha W Mahungu ^91^.

##### Sequencing, analysis, project administration and metadata curation

Rebecca Dewar ^20^, and Martin P McHugh ^20^.

##### Samples, logistics, metadata curation and project administration

Natasha G Jesudason ^21^, Kathy K Li MBBCh ^21^, Rajiv N Shah ^21^, and Yusri Taha ^66^.

##### Leadership, supervision, funding acquisition and metadata curation

Kate E Templeton ^20^.

##### Leadership, supervision, funding acquisition, sequencing and analysis

Simon Cottrell ^33^, Justin O’Grady ^51^, Andrew Rambaut ^19^, and Colin P Smith^93^.

##### Leadership, supervision, metadata curation, sequencing and analysis

Matthew T.G. Holden ^87^, and Emma C Thomson ^21^.

##### Leadership, supervision, samples, logistics and metadata curation

Samuel Moses ^81, 82^.

##### Sequencing, analysis, leadership, supervision, samples and logistics

Meera Chand ^7^, Chrystala Constantinidou ^71^, Alistair C Darby ^46^, Julian A Hiscox ^46^, Steve Paterson ^46^, and Meera Unnikrishnan ^71^.

##### Sequencing, analysis, leadership and supervision and software and analysis tools

Andrew J Page ^51^, and Erik M Volz ^96^.

##### Samples, logistics, sequencing, analysis and metadata curation

Charlotte J Houldcroft ^8^, Aminu S Jahun ^11^, James P McKenna ^88^, Luke W Meredith ^11^, Andrew Nelson ^61^, Sarojini Pandey ^72^, and Gregory R Young ^60^.

##### Sequencing, analysis, metadata curation, and software and analysis tools

Anna Price ^34^, Sara Rey ^33^, Sunando Roy ^41^, Ben Temperton^49^, and Matthew Wyles ^38^.

##### Sequencing, analysis, metadata curation and visualisation

Stefan Rooke^19^, and Sharif Shaaban ^87^.

##### Visualisation, sequencing, analysis and software and analysis tools

Helen Adams ^35^, Yann Bourgeois ^69^, Katie F Loveson ^68^, Áine O’Toole ^19^, and Richard Stark ^71^.

##### Project administration, leadership and supervision

Ewan M Harrison ^1, 3^, David Heyburn ^33^, and Sharon J Peacock ^2, 3^

##### Project administration and funding acquisition

David Buck ^26^, and Michaela John^36^

##### Sequencing, analysis and project administration

Dorota Jamrozy ^1^, and Joshua Quick ^15^

##### Samples, logistics, and project administration

Rahul Batra ^78^, Katherine L Bellis ^1, 3^, Beth Blane ^3^, Sophia T Girgis ^3^, Angie Green ^26^, Anita Justice ^28^, Mark Kristiansen ^41^, and Rachel J Williams ^41^.

##### Project administration, software and analysis tools

Radoslaw Poplawski^15^.

##### Project administration and visualisation

Garry P Scarlett ^69^.

##### Leadership, supervision, and funding acquisition

John A Todd ^26^, Christophe Fraser ^27^, Judith Breuer ^40,41^, Sergi Castellano ^41^, Stephen L Michell^49^, Dimitris Gramatopoulos ^73^, and Jonathan Edgeworth ^78^.

##### Leadership, supervision and metadata curation

Gemma L Kay ^51^.

##### Leadership, supervision, sequencing and analysis

Ana da Silva Filipe ^21^, Aaron R Jeffries ^49^, Sascha Ott ^71^, Oliver Pybus ^24^, David L Robertson ^21^, David A Simpson ^6^, and Chris Williams ^33^.

##### Samples, logistics, leadership and supervision

Cressida Auckland ^50^, John Boyes ^83^, Samir Dervisevic ^52^, Sian Ellard ^49, 50^, Sonia Goncalves^1^, Emma J Meader ^51^, Peter Muir ^2^, Husam Osman ^95^, Reenesh Prakash ^52^, Venkat Sivaprakasam

^18^, and Ian B Vipond ^2^.

##### Leadership, supervision and visualisation

Jane AH Masoli ^49, 50^.

##### Sequencing, analysis and metadata curation

Nabil-Fareed Alikhan ^51^, Matthew Carlile ^54^, Noel Craine ^33^, Sam T Haldenby ^46^, Nadine Holmes ^54^, Ronan A Lyons ^37^, Christopher Moore ^54^, Malorie Perry ^33^, Ben Warne ^80^, and Thomas Williams ^19^.

##### Samples, logistics and metadata curation

Lisa Berry ^72^, Andrew Bosworth ^95^, Julianne Rose Brown ^40^, Sharon Campbell ^67^, Anna Casey^17^, Gemma Clark ^56^, Jennifer Collins ^66^, Alison Cox ^43, 44^, Thomas Davis ^84^, Gary Eltringham ^66^,Cariad Evans ^38, 39^, Clive Graham ^64^, Fenella Halstead ^18^, Kathryn Ann Harris ^40^, Christopher Holmes ^58^, Stephanie Hutchings ^2^, Miren Iturriza-Gomara ^46^, Kate Johnson ^38, 39^, Katie Jones ^72^, Alexander J Keeley ^38^, Bridget A Knight ^49, 50^, Cherian Koshy^90^, Steven Liggett ^63^, Hannah Lowe ^81^, Anita O Lucaci ^46^, Jessica Lynch ^25, 29^, Patrick C McClure ^55^, Nathan Moore ^31^, Matilde Mori ^25, 29, 32^, David G Partridge ^38, 39^, Pinglawathee Madona ^43, 44^, Hannah MPymont ^2^, Paul Anthony Randell ^43, 44^, Mohammad Raza ^38, 39^, Felicity Ryan ^81^, Robert Shaw^28^, Tim J Sloan ^57^, and Emma Swindells ^65^.

##### Sequencing, analysis, Samples and logistics

Alexander Adams ^33^, Hibo Asad ^33^, Alec Birchley ^33^, Tony Thomas Brooks ^41^, Giselda Bucca ^93^, Ethan Butcher ^70^, Sarah L Caddy ^13^, Laura G Caller ^2, 3, 12^, Yasmin Chaudhry ^11^, Jason Coombes^33^, Michelle Cronin ^33^, Patricia L Dyal ^41^, Johnathan M Evans ^33^, Laia Fina ^33^, Bree Gatica-Wilcox ^33^, Iliana Georgana ^11^, Lauren Gilbert ^33^, Lee Graham ^33^, Danielle C Groves ^38^, Grant Hall ^11^, Ember Hilvers ^33^, Myra Hosmillo ^11^, Hannah Jones ^33^, Sophie Jones ^33^, Fahad A Khokhar ^13^, Sara Kumziene-Summerhayes ^33^, George MacIntyre-Cockett ^26^, Rocio T Martinez Nunez ^94^, Caoimhe McKerr ^33^, Claire McMurray ^15^, Richard Myers ^7^, Yasmin Nicole Panchbhaya ^41^, Malte L Pinckert ^11^, Amy Plimmer ^33^, Joanne Stockton ^15^, Sarah Taylor ^33^, Alicia Thornton ^7^, Amy Trebes ^26^, Alexander J Trotter ^51^, Helena Jane Tutill ^41^, Charlotte A Williams ^41^, Anna Yakovleva ^11^ and Wen C Yew ^62^.

##### Sequencing, analysis and software and analysis tools

Mohammad T Alam ^71^, Laura Baxter ^71^, Olivia Boyd ^96^, Fabricia F. Nascimento ^96^, Timothy M Freeman ^38^, Lily Geidelberg ^96^, Joseph Hughes ^21^, David Jorgensen ^96^, Benjamin B Lindsey ^38^,Richard J Orton ^21^, Manon Ragonnet-Cronin ^96^ Joel Southgate ^33, 34^, and Sreenu Vattipally ^21^.

##### Samples, logistics and software and analysis tools

Igor Starinskij ^23^.

##### Visualisation and software and analysis tools

Joshua B Singer ^21^, Khalil Abudahab ^1, 30^, Leonardo de Oliveira Martins ^51^, Thanh Le-Viet ^51^, Mirko Menegazzo ^30^, Ben EW Taylor ^1, 30^, and Corin A Yeats ^30^.

##### Project Administration

Sophie Palmer ^3^, Carol M Churcher ^3^, Alisha Davies ^33^, Elen De Lacy ^33^, Fatima Downing ^33^, Sue Edwards ^33^, Nikki Smith ^38^, Francesc Coll ^97^, Nazreen F Hadjirin ^3^ and Frances Bolt ^44, 45^.

##### Leadership and supervision

Alex Alderton^1^, Matt Berriman^1^, Ian G Charles ^51^, Nicholas Cortes ^31^, Tanya Curran ^88^, John Danesh^1^, Sahar Eldirdiri ^84^, Ngozi Elumogo ^52^, Andrew Hattersley ^49, 50^, Alison Holmes ^44, 45^, Robin Howe ^33^, Rachel Jones ^33^, Anita Kenyon ^84^, Robert A Kingsley ^51^, Dominic Kwiatkowski ^1, 9^, Cordelia Langford^1^, Jenifer Mason^48^, Alison E Mather ^51^, Lizzie Meadows ^51^, Sian Morgan ^36^, James Price ^44, 45^, Trevor I Robinson ^48^, Giri Shankar ^33^, John Wain ^51^, and Mark A Webber ^51^.

##### Metadata curation

Declan T Bradley ^5, 6^, Michael R Chapman ^1, 3, 4^, Derrick Crooke ^28^, David Eyre ^28^, Martyn Guest^34^, Huw Gulliver ^34^, Sarah Hoosdally ^28^, Christine Kitchen ^34^, Ian Merrick ^34^, SiddharthMookerjee ^44, 45^, Robert Munn ^34^, Timothy Peto ^28^, Will Potter ^52^, Dheeraj K Sethi ^52^, Wendy Smith ^56^, Luke B Snell ^75, 94^, Rachael Stanley ^52^, Claire Stuart ^52^ and Elizabeth Wastenge^20^.

##### Sequencing and analysis

Erwan Acheson ^6^, Safiah Afifi ^36^, Elias Allara ^2, 3^, Roberto Amato ^1^, Adrienn Angyal ^38^, Elihu Aranday-Cortes ^21^, Cristina Ariani ^1^, Jordan Ashworth ^19^, Stephen Attwood ^24^, Alp Aydin ^51^, David J Baker ^51^, Carlos E Balcazar ^19^, Angela Beckett ^68^ Robert Beer ^36^, Gilberto Betancor ^76^, Emma Betteridge ^1^, David Bibby ^7^, Daniel Bradshaw^7^, Catherine Bresner ^34^, Hannah E Bridgewater ^71^, Alice Broos ^21^, Rebecca Brown ^38^, Paul E Brown ^71^, Kirstyn Brunker ^22^, Stephen N Carmichael ^21^, Jeffrey K. J. Cheng ^71^, Dr Rachel Colquhoun ^19^, Gavin Dabrera ^7^, Johnny Debebe ^54^, Eleanor Drury ^1^, Louis du Plessis ^24^, Richard Eccles ^46^, Nicholas Ellaby ^7^, Audrey Farbos ^49^, Ben Farr ^1^, Jacqueline Findlay ^41^, Chloe L Fisher ^74^, Leysa Marie Forrest ^41^, Sarah Francois ^24^, Lucy R. Frost ^71^, William Fuller^34^, Eileen Gallagher ^7^, Michael D Gallagher ^19^, Matthew Gemmell ^46^, Rachel AJ Gilroy ^51^, Scott Goodwin ^1^, Luke R Green ^38^, Richard Gregory ^46^, Natalie Groves ^7^, James W Harrison ^49^, Hassan Hartman ^7^, Andrew R Hesketh ^93^,Verity Hill ^19^, Jonathan Hubb ^7^, Margaret Hughes^46^, David K Jackson ^1^, Ben Jackson ^19^, Keith James ^1^,Natasha Johnson ^21^, Ian Johnston ^1^, Jon-Paul Keatley ^1^, Moritz Kraemer ^24^, Angie Lackenby ^7^, Mara Lawniczak ^1^, David Lee ^7^, Rich Livett ^1^, Stephanie Lo ^1^, Daniel Mair ^21^, Joshua Maksimovic ^36^, Nikos Manesis ^7^, Robin Manley ^49^, Carmen Manso ^7^, Angela Marchbank ^34^, Inigo Martincorena ^1^, Tamyo Mbisa ^7^, Kathryn McCluggage ^36^, JT McCrone ^19^, Shahjahan Miah ^7^, Michelle L Michelsen ^49^, Mari Morgan ^33^, Gaia Nebbia ^78^,Charlotte Nelson ^46^, Jenna Nichols ^21^, Paola Niola ^41^, Kyriaki Nomikou ^21^, Steve Palmer ^1^, Naomi Park ^1^, Yasmin A Parr ^1^, Paul J Parsons ^38^, Vineet Patel ^7^, Minal Patel ^1^, Clare Pearson ^2, 1^, Steven Platt ^7^, Christoph Puethe ^1^, Mike Quail ^1^,Jayna Raghwani ^24^, Lucille Rainbow ^46^, Shavanthi Rajatileka ^1^, Mary Ramsay ^7^, Paola C Resende Silva ^41, 42^, Steven Rudder 51, Chris Ruis ^3^, Christine M Sambles ^49^, Fei Sang ^54^, Ulf Schaefer^7^, Emily Scher ^19^, Carol Scott ^1^, Lesley Shirley ^1^, Adrian W Signell ^76^, John Sillitoe ^1^, Christen Smith ^1^, Dr Katherine L Smollett ^21^, Karla Spellman ^36^, Thomas D Stanton ^19^, David J Studholme ^49^, Grace Taylor-Joyce ^71^, Ana P Tedim ^51^, Thomas Thompson ^6^,Nicholas M Thomson ^51^, Scott Thurston^1^, Lily Tong ^21^, Gerry Tonkin-Hill ^1^, Rachel M Tucker ^38^, Edith E Vamos ^4^, Tetyana Vasylyeva^24^, Joanna Warwick-Dugdale ^49^, Danni Weldon ^1^, Mark Whitehead ^46^, David Williams ^7^, Kathleen A Williamson ^19^,Harry D Wilson ^76^,Trudy Workman^34^, Muhammad Yasir^51^, Xiaoyu Yu ^19^, and Alex Zarebski ^24^.

##### Samples and logistics

Evelien M Adriaenssens ^51^, Shazaad S Y Ahmad ^2, 47^, Adela Alcolea-Medina ^59, 77^, John Allan ^60^, Patawee Asamaphan ^21^, Laura Atkinson ^40^, Paul Baker ^63^, Jonathan Ball ^55^, Edward Barton^64^, Mathew A Beale^1^, Charlotte Beaver^1^, Andrew Beggs ^16^, Andrew Bell ^51^, Duncan J Berger ^1^, Louise Berry. ^56^, Claire M Bewshea ^49^, Kelly Bicknell ^70^, Paul Bird ^58^, Chloe Bishop ^7^, Tim Boswell ^56^, Cassie Breen ^48^, Sarah K Buddenborg^1^, Shirelle Burton-Fanning ^66^, Vicki Chalker ^7^, Joseph G Chappell ^55^, Themoula Charalampous ^78, 94^, Claire Cormie^3^, Nick Cortes^29, 25^, Lindsay J Coupland ^52^, Angela Cowell ^48^, Rose K Davidson ^53^, Joana Dias ^3^, Maria Diaz ^51^, Thomas Dibling^1^, Matthew J Dorman^1^, Nichola Duckworth^57^, Scott Elliott^70^, Sarah Essex^63^, Karlie Fallon ^58^, Theresa Feltwell ^8^, Vicki M Fleming ^56^, Sally Forrest ^3^, Luke Foulser^1^, Maria V Garcia-Casado^1^, Artemis Gavriil ^41^, Ryan P George ^47^, Laura Gifford ^33^, Harmeet K Gill ^3^, Jane Greenaway ^65^, Luke Griffith^53^, Ana Victoria Gutierrez^51^, Antony D Hale ^85^, Tanzina Haque ^91^,Katherine L Harper ^85^, Ian Harrison ^7^, Judith Heaney ^89^, Thomas Helmer ^58^, Ellen E Higginson^3^, Richard Hopes ^2^, Hannah C Howson-Wells ^56^, Adam D Hunter ^1^, Robert Impey ^70^, Dianne Irish-Tavares ^91^, David A Jackson^1^, Kathryn A Jackson ^46^, Amelia Joseph ^56^, Leanne Kane ^1^,Sally Kay ^1^, Leanne M Kermack ^3^, Manjinder Khakh ^56^, Stephen P Kidd ^29, 25,31^, Anastasia Kolyva^51^, Jack CD Lee ^40^, Laura Letchford ^1^, Nick Levene ^79^, Lisa J Levett ^89^, Michelle M Lister ^56^,Allyson Lloyd ^70^, Joshua Loh ^60^, Louissa R Macfarlane-Smith ^85^, Nicholas W Machin ^2, 47^, Mailis Maes ^3^, Samantha McGuigan ^1^, Liz McMinn ^1^, Lamia Mestek-Boukhibar ^41^, Zoltan Molnar ^6^, Lynn Monaghan ^79^, Catrin Moore ^27^, Plamena Naydenova ^3^, Alexandra S Neaverson ^1^, Rachel Nelson ^1^, Marc O Niebel ^21^, Elaine O’Toole^48^, Debra Padgett ^64^, Gaurang Patel ^1^, Brendan AI Payne ^66^, Liam Prestwood ^1^, Veena Raviprakash ^67^, Nicola Reynolds^86^, Alex Richter ^16^, Esther Robinson ^95^, Hazel A Rogers^1^, Aileen Rowan ^96^, Garren Scott ^64^, Divya Shah ^40^, Nicola Sheriff ^67^, Graciela Sluga, Emily Souster^1^, Michael Spencer-Chapman^1^, Sushmita Sridhar ^1, 3^, Tracey Swingler ^53^, Julian Tang^58^, Graham P Taylor^96^, Theocharis Tsoleridis ^55^, Lance Turtle^46^, Sarah Walsh ^57^, Michelle Wantoch ^86^, Joanne Watts ^48^, Sheila Waugh ^66^, Sam Weeks^41^, Rebecca Williams^31^, Iona Willingham^56^, Emma L Wise ^25, 29, 31^, Victoria Wright ^54^, Sarah Wyllie ^70^, and Jamie Young ^3^.

##### Software and analysis tools

Amy Gaskin^33^, Will Rowe ^15^, and Igor Siveroni ^96^.

##### Visualisation

Robert Johnson ^96^.

**1** Wellcome Sanger Institute, **2** Public Health England, **3** University of Cambridge, **4** Health Data Research UK, Cambridge, **5** Public Health Agency, Northern Ireland, **6** Queen’s University Belfast **7** Public Health England Colindale, **8** Department of Medicine, University of Cambridge, **9** University of Oxford, **10** Departments of Infectious Diseases and Microbiology, Cambridge University Hospitals NHS Foundation Trust; Cambridge, UK, **11** Division of Virology, Department of Pathology, University of Cambridge, **12** The Francis Crick Institute, **13** Cambridge Institute for Therapeutic Immunology and Infectious Disease, Department of Medicine, **14** Public Health England, Clinical Microbiology and Public Health Laboratory, Cambridge, UK, **15** Institute of Microbiology and Infection, University of Birmingham, **16** University of Birmingham, **17** Queen Elizabeth Hospital, **18** Heartlands Hospital, **19** University of Edinburgh, **20** NHS Lothian, **21** MRC-University of Glasgow Centre for Virus Research, **22** Institute of Biodiversity, Animal Health & Comparative Medicine, University of Glasgow, **23** West of Scotland Specialist Virology Centre, **24** Dept Zoology, University of Oxford, **25** University of Surrey, **26** Wellcome Centre for Human Genetics, Nuffield Department of Medicine, University of Oxford, **27** Big Data Institute, Nuffield Department of Medicine, University of Oxford, **28** Oxford University Hospitals NHS Foundation Trust, **29** Basingstoke Hospital, **30** Centre for Genomic Pathogen Surveillance, University of Oxford, **31** Hampshire Hospitals NHS Foundation Trust, **32** University of Southampton, **33** Public Health Wales NHS Trust, **34** Cardiff University, **35** Betsi Cadwaladr University Health Board, **36** Cardiff and Vale University Health Board, **37** Swansea University, **38** University of Sheffield, **39** Sheffield Teaching Hospitals, **40** Great Ormond Street NHS Foundation Trust, **41** University College London, **42** Oswaldo Cruz Institute, Rio de Janeiro **43** North West London Pathology, **44** Imperial College Healthcare NHS Trust, **45** NIHR Health Protection Research Unit in HCAI and AMR, Imperial College London, **46** University of Liverpool, **47** Manchester University NHS Foundation Trust, **48** Liverpool Clinical Laboratories, **49** University of Exeter, **50** Royal Devon and Exeter NHS Foundation Trust, **51** Quadram Institute Bioscience, University of East Anglia, **52** Norfolk and Norwich University Hospital, **53** University of East Anglia, **54** Deep Seq, School of Life Sciences, Queens Medical Centre, University of Nottingham, **55** Virology, School of Life Sciences, Queens Medical Centre, University of Nottingham, **56** Clinical Microbiology Department, Queens Medical Centre, **57** PathLinks, Northern Lincolnshire & Goole NHS Foundation Trust, **58** Clinical Microbiology, University Hospitals of Leicester NHS Trust, **59** Viapath, **60** Hub for Biotechnology in the Built Environment, Northumbria University, **61** NU-OMICS Northumbria University, **62** Northumbria University, **63** South Tees Hospitals NHS Foundation Trust, **64** North Cumbria Integrated Care NHS Foundation Trust, **65** North Tees and Hartlepool NHS Foundation Trust, **66** Newcastle Hospitals NHS Foundation Trust, **67** County Durham and Darlington NHS Foundation Trust, **68** Centre for Enzyme Innovation, University of Portsmouth, **69** School of Biological Sciences, University of Portsmouth, **70** Portsmouth Hospitals NHS Trust, **71** University of Warwick, **72** University Hospitals Coventry and Warwickshire, **73** Warwick Medical School and Institute of Precision Diagnostics, Pathology, UHCW NHS Trust, **74** Genomics Innovation Unit, Guy’s and St. Thomas’ NHS Foundation Trust, **75** Centre for Clinical Infection & Diagnostics Research, St. Thomas’ Hospital and Kings College London, **76** Department of Infectious Diseases, King’s College London, **77** Guy’s and St. Thomas’ Hospitals NHS Foundation Trust, **78** Centre for Clinical Infection and Diagnostics Research, Department of Infectious Diseases, Guy’s and St Thomas’ NHS Foundation Trust, **79** Princess Alexandra Hospital Microbiology Dept., **80** Cambridge University Hospitals NHS Foundation Trust, **81** East Kent Hospitals University NHS Foundation Trust, **82** University of Kent, **83** Gloucestershire Hospitals NHS Foundation Trust, **84** Department of Microbiology, Kettering General Hospital, **85** National Infection Service, PHE and Leeds Teaching Hospitals Trust, **86** Cambridge Stem Cell Institute, University of Cambridge, **87** Public Health Scotland, 88 Belfast Health & Social Care Trust, **89** Health Services Laboratories, **90** Barking, Havering and Redbridge University Hospitals NHS Trust, **91** Royal Free NHS Trust, **92** Maidstone and Tunbridge Wells NHS Trust, **93** University of Brighton, **94** Kings College London, **95** PHE Heartlands, **96** Imperial College London, **97** Department of Infection Biology, London School of Hygiene and Tropical Medicine.

